# Retinograd-AI: An Open-source Automated Fundus Autofluorescence Retinal Image Gradability Assessment for Inherited Retinal Dystrophies

**DOI:** 10.1101/2024.08.07.24311607

**Authors:** Gunjan Naik, Saoud Al-Khuzaei, Ismail Moghul, Thales A. C. de Guimaraes, Sagnik Sen, Malena Daich Varela, Yichen Liu, Pallavi Bagga, Dun Jack Fu, Mariya Moosajee, Savita Madhusudhan, Andrew Webster, Samantha De Silva, Praveen J. Patel, Omar Mahroo, Susan M Downes, Michel Michaelides, Konstantinos Balaskas, Nikolas Pontikos, William Woof

## Abstract

**Purpose:** To develop an automated system for assessing the quality of Fundus Autofluorescence (FAF) images in patients with inherited retinal diseases (IRD).

**Methods:** We annotated a dataset of 2445 FAF images from patients with Inherited Retinal Dystrophies which were assessed by three different expert graders. Graders marked images as either gradable (acceptable quality) or ungradable (poor quality), following a strict grading protocol. This dataset was used to train a Convolutional Neural Network (CNN) classification model to predict the gradability label of FAF images.

**Results:** Retinograd-AI achieves a performance of 91% accuracy on our held-out dataset of 133 images with an Area Under the Receiver Operator Characteristic (AUROC) of 0.94, indicating high performance in distinguishing between gradable and ungradable images. Applying Retinograd-AI to our full internal dataset, the highest proportion of gradable images was found in the 30-50 years age group, where 84.3% of images were rated as gradable, while the lowest was in 0-15 year olds, where only 45.2% of images were rated as gradable. 83.4% of images from male patients were rated as gradable, and 90.6% of images from female patients. By genotype, from the 30 most common genetic diagnoses, the highest proportion of gradable images was in patients with disease causing variants in *PRPH2* (93.9%), while the lowest was *RDH12* (28.6%). Eye2Gene single-image gene classification top-5 accuracy on images rated by Retinograd-AI was 69.2%, while top-5 accuracy on images rated as ungradable was 39.0%. Retinograd-AI is open-sourced, and the source code and network weights are available under an MIT licence on GitHub at https://github.com/Eye2Gene/retinograd-ai

**Conclusions:** Retinograd-AI is the first open-source AI model for automated retinal image quality assessment of FAF images in IRDs. Automated gradability assessment through Retinograd AI enables large scale analysis of retinal images, which is an essential part of developing good analysis pipelines, and real-time quality assessment, which is essential for deployment of AI algorithms, such as Eye2Gene, into clinical settings. Due to the diverse nature of IRD pathologies, Retinograd-AI may also be applicable to FAF imaging for other conditions, either in its current form or through transfer learning and fine-tuning.

## Introduction

Inherited Retinal Dystrophies (IRDs) are genetically determined disorders of the retina, which collectively represent a leading cause of blindness in children and the working-age population. IRDs encompass a wide range of conditions with 280 different associated genes identified so far (Georgiou et al., 2024; Lee et al., 2023; Pontikos et al., 2020). Retinal imaging, using various imaging modalities, allows accurate phenotyping, which is important in the diagnosis and follow-up of IRDs.

Fundus autofluorescence (FAF) imaging is particularly important in this regard since it can yield data relating to the outer retina and retinal pigment epithelium (RPE). For instance, an increased autofluorescent signal (hyper autofluorescence) can result from the accumulation of autofluorescent material, such as lipofuscin, or from the loss of either photoreceptor outer segments or macular luteal pigment, which usually absorbs the incoming short wavelengths (Daich Varela et al., 2021). Similarly, loss of autofluorescence can be associated with the loss of RPE. Particular patterns of autofluorescence are associated with certain IRDs, such as the hyper autofluorescent flecks that are usually associated with Stargardt disease (Pichi et al., 2018).

The quality of imaging data is a critical factor for developing AI models and in particular during selection of scans for training AI models such as Eye2Gene and AIRDetect (Nguyen et al., 2023; Pontikos et al., 2022; Woof et al., 2024). Image quality significantly influences model performance and uncertainty metrics in image classification or segmentation. Poor quality images frequently cause AI model failures, whereas clinicians would disregard these as ungradable or request repeat imaging.

Image gradeability refers to if an image is sufficient for a human (or AI) specialist to make an informed decision on the basis of the image. Although gradability is technically distinct from image quality, these aspects are highly correlated and in the literature the terms are often used interchangeably (Huynh et al., 2024). Manually grading images is laborious and subjective, which highlights the need for automated gradability assessment to filter out poor-quality imaging data. This is crucial for selecting images for training AI models and for using these models to assess biomarkers in clinical trials. These approaches are also necessary for deployment of AI systems in the real world setting by employing automated grading as a pre-filtering step to assess whether repeat imaging is necessary and prevent propagation of decisions based on unreliable data.

Several AI models for retinal image quality assessment of colour fundus images have been developed (Abdel-Hamid, 2021; Abramovich et al., 2023; Chan et al., 2021; Shi et al., 2022; J. Tang et al., 2022), as well as a few for assessing the quality of Spectral domain OCT (SD-OCT) images (Z. Tang et al., 2024; Wang et al., 2019). However, no models currently exist for other modalities such as fundus autofluorescence (FAF), and none have been specifically developed for the gradability of IRDs.

Assessment of gradability of retinal scans from IRD patients poses unique challenges due IRDs having a range of phenotypes depending on the gene involved. For example, large areas of abnormal retina can often obstruct features such as the retinal vasculature, or decreased autofluorescence can render regions darker than usual. Distinguishing these pathological features from other imaging artefacts is challenging, but is crucial for reliable grading and the proper functioning of AI models. Hence, in addition to their application to IRDs, IRD datasets may be a good starting point for developing more general gradability assessment models, as they encompass a wide range of different conditions and pathologies, and affect patients across all age ranges.

We present Retinograd-AI, the first retinal image gradability assessment tool for FAF imaging and the first specifically developed for all types of IRDs. Retinograd-AI is a deep neural network (DNN) based classifier trained and validated on over 2400 FAF images from patients seen at Moorfields Eye Hospital (MEH), annotated by three expert graders.

## Methods

### Dataset

Our training dataset was drawn from a dataset of a total of 136,631 FAF images from 4,554 IRD patients from Moorfields Eye Hospital (MEH), captured using the Heidelberg Spectralis imaging platform. From this dataset, 2445 images were selected at random and then labelled by a team of three graders (G1, G2, G3), with 815 images assigned to each grader. All graders were research fellows with over 5 years’ experience in medical retina, and had extensive experience with grading FAF scans for IRDs. Annotation was performed using two defined criteria for image quality, as outlined in **Table 1**. This annotation was done over the course of three weeks using an instance of the Label Studio tool (Tkachenko et al., 2020-2022), which was hosted on our online grading platform (grading.readingcentre.org).

**Table 1:** Definition of the quality assessment criteria.

| Assessment | Criteria | Feature Visibility |
| --- | --- | --- |
| **Gradable**<br>**(acceptable**<br>**quality to a**<br>**grader)** | Image is sufficient to yield a grade with >50% certainty | Discrimination of the optic disc is clear and vascular arcades are visible in over ¾ of their extent.<br>No opacities/shadowing impairing clear visibility of critical structures like the foveal and peri-foveal areas. |
| **Ungradable**<br>**(un-acceptable**<br>**quality to a**<br>**grader)** | Image is not sufficient to yield a grade. | One or more anatomical features impossible to discern. |

An additional 133 images were selected as a held-out test set, ensuring no patient overlap with the training set, each of which were annotated by all three graders. This was used to measure intergrader agreement and evaluate our algorithm. In cases where not all graders agreed on the same label for a given image, the most common label was used for the purposes of model evaluation. This approach helped ensure consistency and reliability in the evaluation process.

### Model Development

For training and evaluation of our Retinograd-AI model, we divided the data into training and pre-test sets in a stratified way, ensuring a similar proportion of each class in both sets by assigning patients to each split to avoid any overlap.

We employed an Inception Resnet v2 network architecture with imagenet pretrained weights for the network. The model was trained using the Adam optimizer and cross-entropy loss, with class reweighting applied to account for dataset imbalance between the two classes.

Horizontal flipping and random rotations to increase variability of data in line with standard data augmentation practices. We have trained the model for 20 epochs, taking the best performing weights determined by the validation loss as evaluated on the pre-test data. A full list of hyper-parameter settings is given in **Supplementary Table 1**.

## Results

The average intergrader agreement was 0.69 as measured by Cohen’s Kappa (McHugh, 2012). A full breakdown of inter-grader agreement is given in **Supplementary Table 2**.

We evaluated Retinograd-AI on the held-out test set and compared its predictions to the grader labels, viewing the problem as a binary classification task with ‘gradable’ being the positive class and ‘ungradable’ being the negative class.

The model achieved an accuracy of 91% (CI_95_=85.7-95.5%) on the held-out test set, with precision of 0.96 (0.923-0.991) and recall of 0.93 (0.873-0.973). The corresponding confusion matrix is given in **Table 2**. The Area Under the Receiver-Operator Characteristic (AUROC) was 0.94 (**Figure 2**). Model-grader agreement (Cohen-Kappa) was 0.69, which was the same as the inter-grader agreement.

**Table 2:** Model confusion matrix. Comparison of model predictions with ground-truth grader labels.

|  |  | Model Prediction |  |
| --- | --- | --- | --- |
|  |  | Gradable | Ungradable |
| Grader Label | Gradable | 104 | 8 |
|  | Ungradable | 4 | 17 |

**Figure 1:**
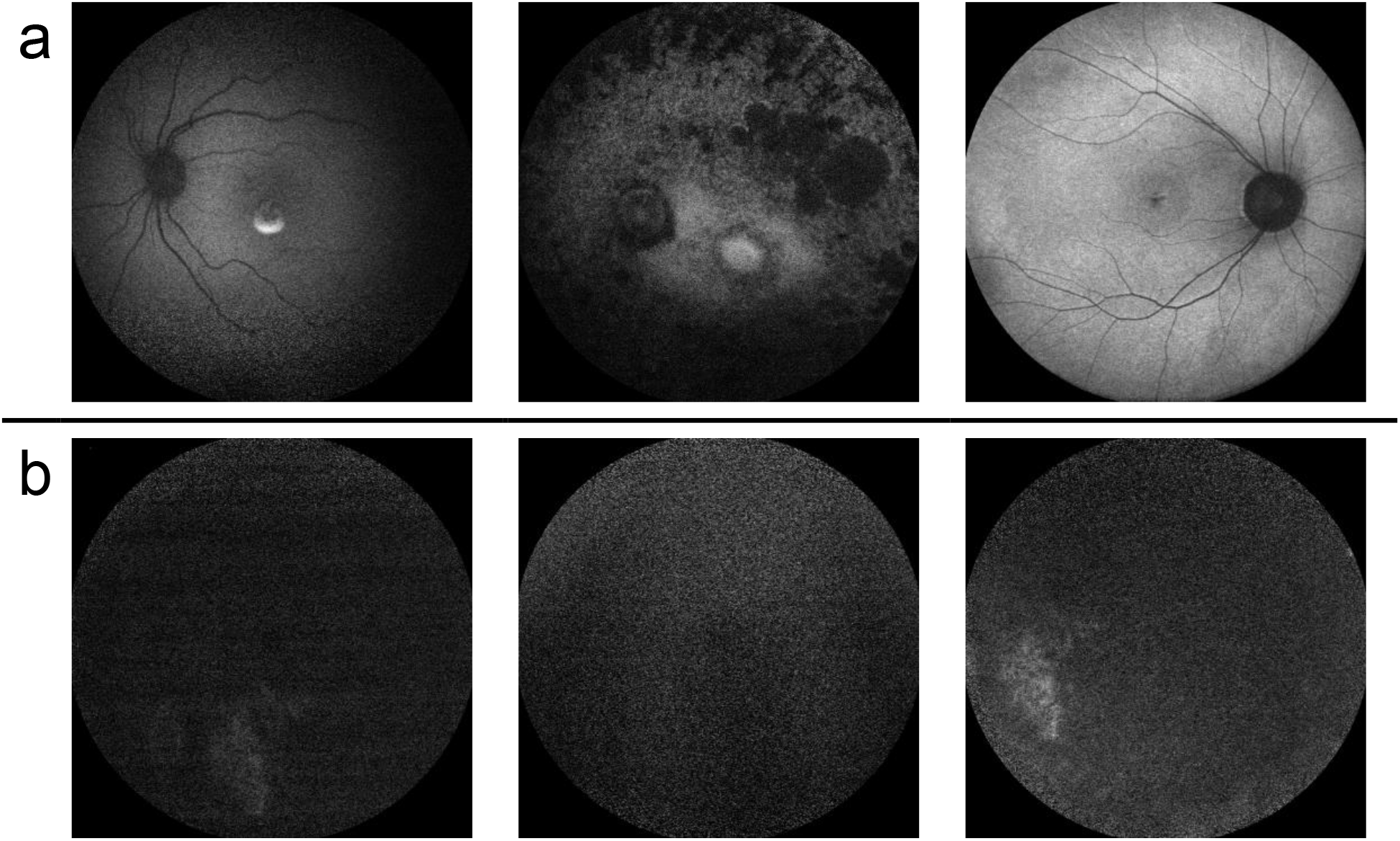
Example images annotated as a.) gradable (acceptable quality), and b.) ungradable (poor quality)

**Figure 2:**
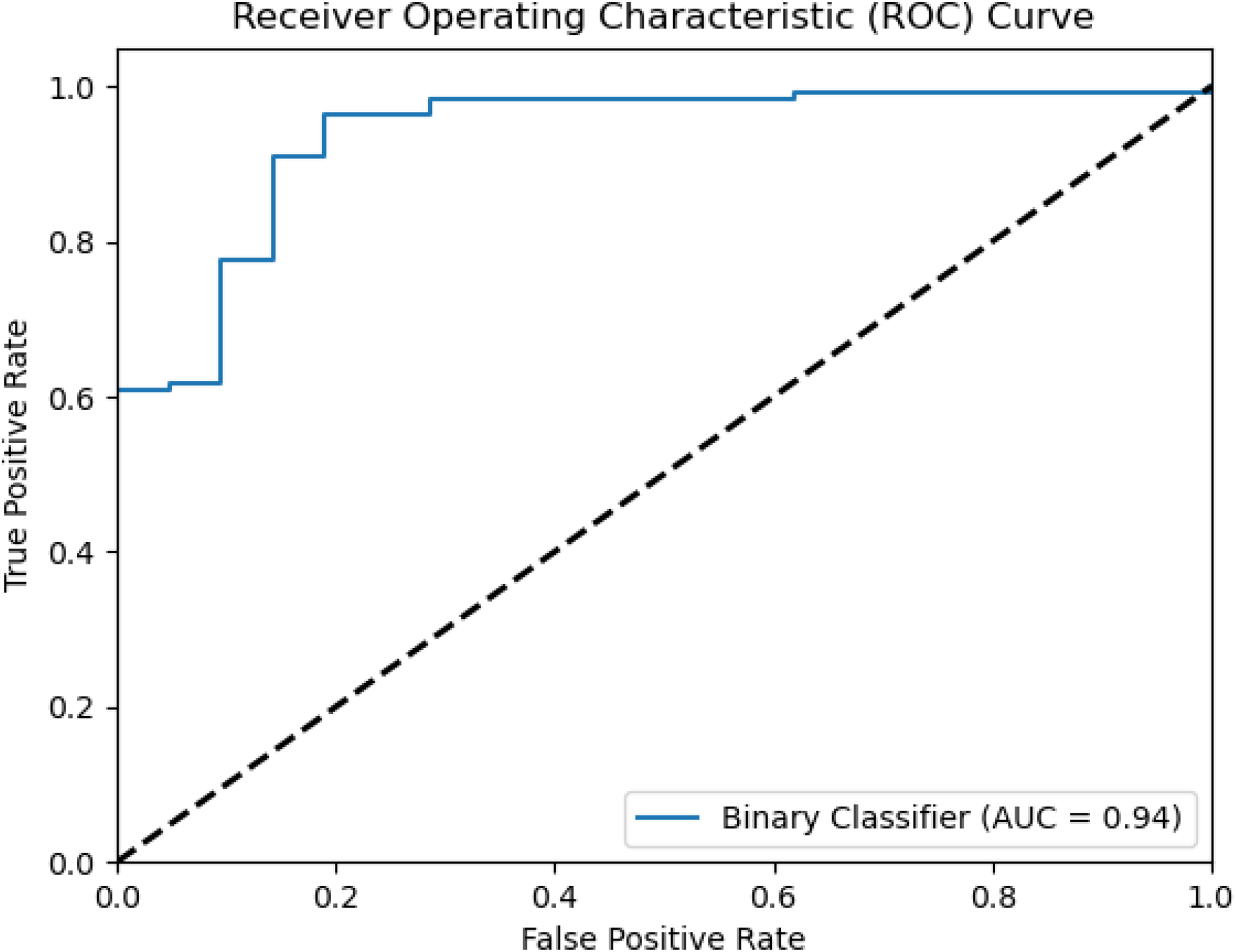
Receiver operator characteristic curve for the model predictions on the held-out test set.

To understand how demographics might affect image quality, we applied Retinograd-AI to our full dataset of 136,631 FAF images, collected as part of the Eye2Gene study, to obtain Retinograd-AI predictions for each image. This enabled us to examine the relationship between image quality and various other data attributes such as patient age and sex, and Eye2Gene classification accuracy.

We observed a mild effect of age on Retinograd-AI assessed image quality, with the highest proportions of gradable images in the 30-50 year old patients, with slightly higher proportion of ungradable images in older patients, and significantly higher proportions in younger patients, particularly in the under-15s (**Table 3)**, which matched expectations. There was also a large difference between male and female patients with images being rated as gradable 83.4% of the time for male patients, and 90.6% for female patients. This difference may be due to the inclusion of X-linked IRDs (*XLRP, CHM*) in the dataset, which affects males more severely than carrier females, leading to more advanced retinal changes and overall poorer image quality in males. There was also substantial variation in image quality across different patient genotypes, with the highest proportion of gradable images were found in patients with a disease-causing variant in *PRPH2*, with 93.9% of images were rated as gradable, and the lowest being *RDH12* where only 28.6% of images were rated as gradable (**Figure 3**).

**Table 3:** Comparison of patient age with gradability.

| Age range | % Gradable |
| --- | --- |
| 0-15 | 45.2% |
| 15-30 | 70.5% |
| 30-50 | 84.3% |
| 50-70 | 82.7% |
| 70+ | 80.7% |

**Figure 3:**
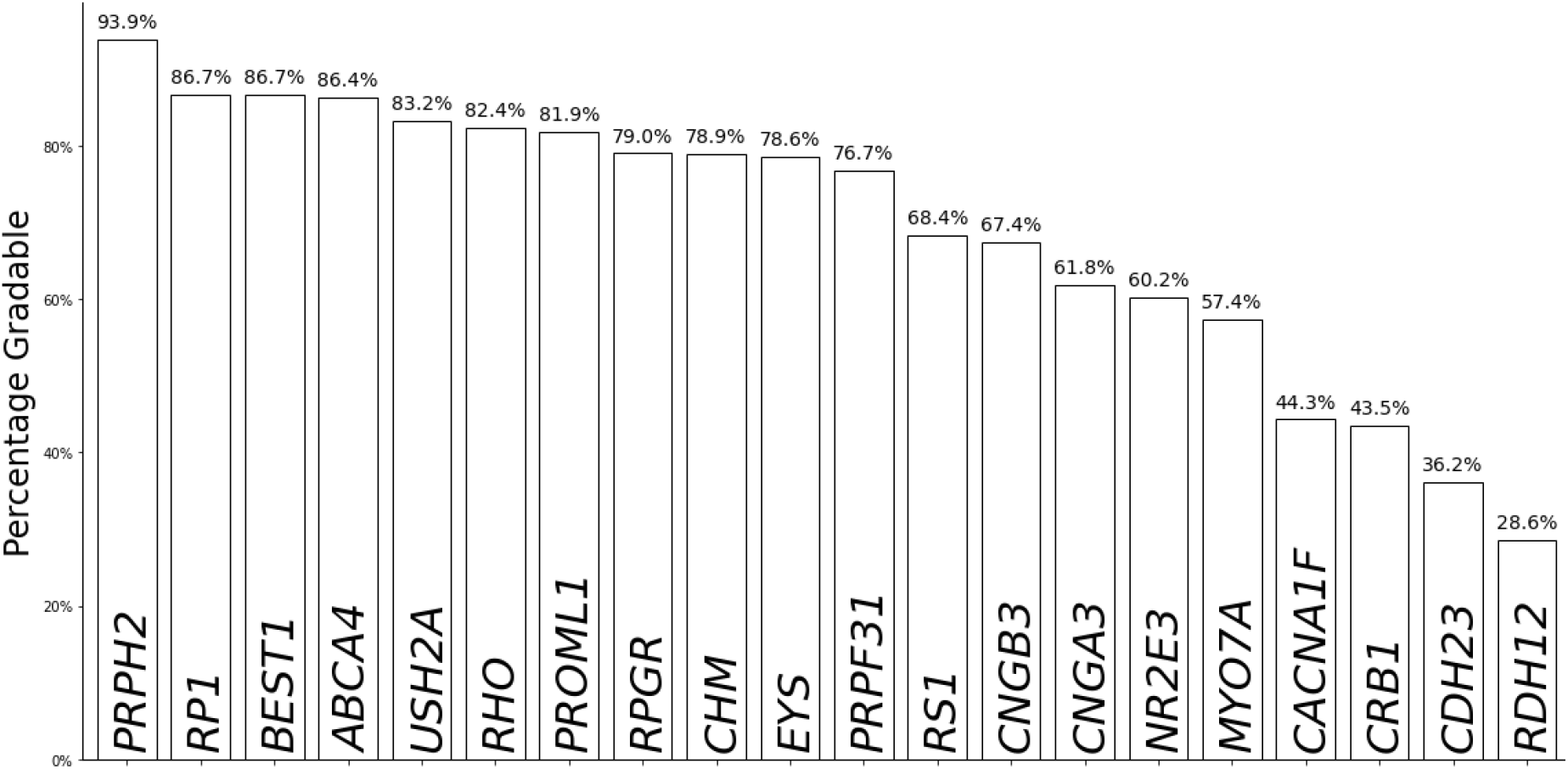
Percentage of images rated as gradable by Retinograd-AI across the 30 most common genetic diagnoses. Significant differences can been seen between genes.

To assess how image quality affects the performance of AI models, we compared the Retinograd-AI assessed image gradeability to the gene-classification accuracy of a single FAF module of Eye2Gene (Nguyen et al., 2023; Pontikos et al., 2022), evaluating at image-level rather than patient-level. We found that images classified as gradable by Retinograd-AI had a top-5 gene classification accuracy of 69.2%, while images classified as ungradable had a substantially lower accuracy of 39.0%. **Figure 4** shows how gene-classification accuracy compares with the raw probability output of Retinograd, showing that higher gradeability score corresponds with higher gene-classification accuracy.

**Figure 4:**
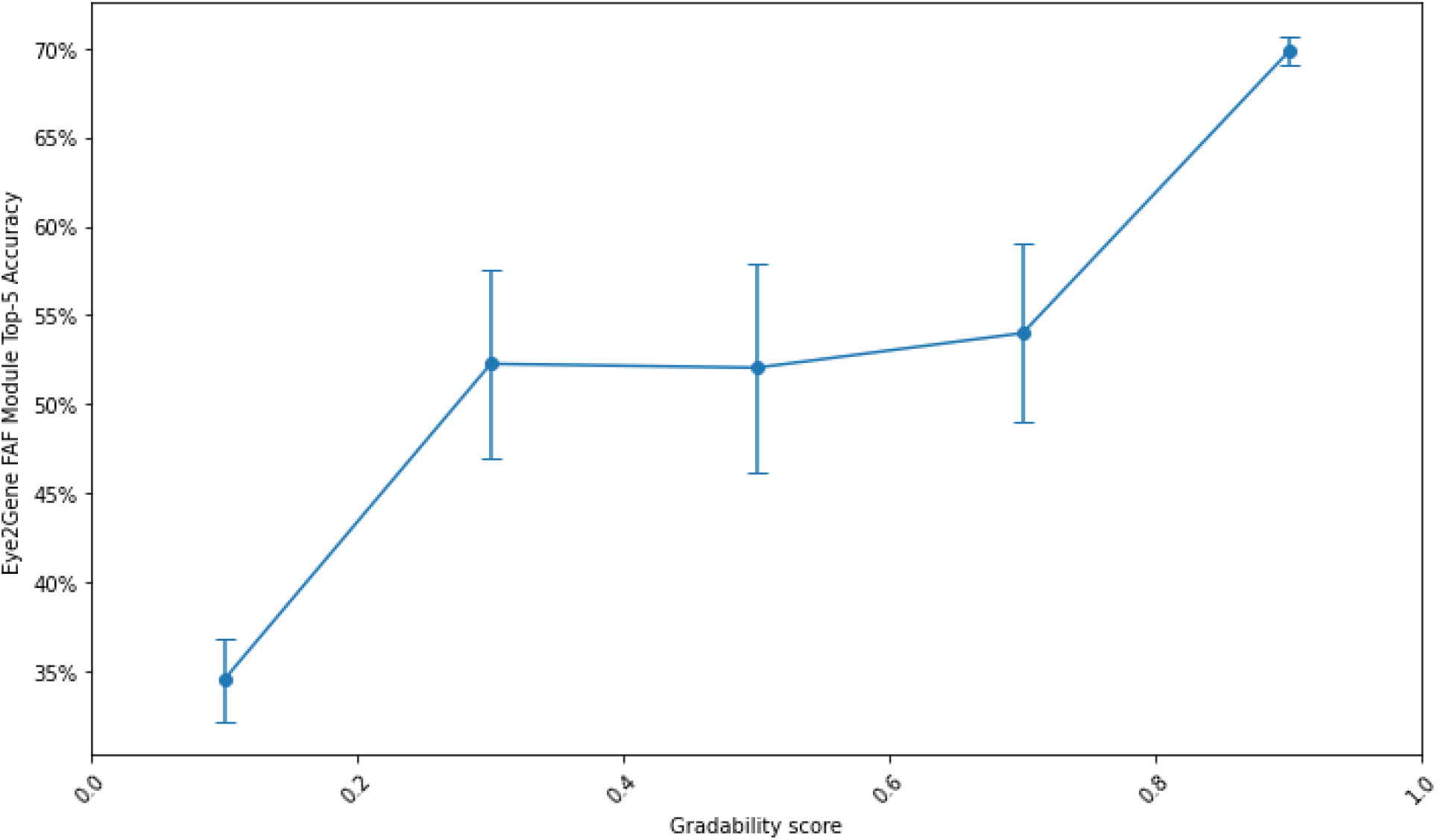
Comparison of Eye2Gene FAF module top-5 gene classification accuracy compared with gradability probability (gradability score) from Retinograd-AI. All images were ranked by the probability output of Retinograd and divided into 5 buckets. For each bucket the per-bucket Eye2Gene classification accuracy, and standard error, were calculated and plotted.

## Discussion

Herein, we present Retinograd-AI, the first retinal image quality assessment model for FAF imaging and the first image quality assessment tool developed specifically for IRDs. We have open-sourced our algorithm to make it available to other researchers at https://github.com/Eye2Gene/retinograd-ai.

Retinograd-AI enabled us to automatically annotate our entire database of FAF images in IRDs from Moorfields Eye Hospital, an otherwise unfeasible task to perform manually. These data enabled us to gain valuable insights into image quality variability in relation to parameters such as patient age, sex and genotype, which historically have been difficult to separate out due to previously-unquantified influences.

As might be expected, we found that younger (0-15 year olds) and older (70+ year olds) age groups had a smaller proportion of gradable images than other age groups. For IRD genotypes, we found that genotypes which are earlier onset, affect the posterior pole such as *RDH12* or cause widespread degeneration such as *CRB1* had a lower proportion of gradable images. As did genotypes that tend to present with secondary cataract, severe phenotypes or high myopia such as *MYO7A, NR2E3* and *CACNA1F*. Achromatopsia genotypes such as *CNGB3* and *CNGBA3* often have nystagmus and photoaversion which could also explain a lower proportion of gradable images in those genotypes.

We were also able to confirm our hypothesis that image quality of FAF imaging has a significant impact on the performance of AI models such as Eye2Gene. This has significant implications for the deployment of AI models into clinical settings.

Changes in the data quality between validation and real-world settings could have a large impact on model performance, leading to substantially lower real-world accuracy than expected, carrying implications for safety and efficacy.

Automated image quality assessment tools, such as Retinograd-AI, can have an important role to play in addressing this, both by identifying variations in image quality between different settings and patient populations, as well as for pre-screening images at point of use to reject poor-quality images.

Retinograd-AI can also be used in other scenarios where image quality is important, but expert feedback is not immediately available, for example, in collecting data for clinical trials.

In these cases Retinograd-AI can provide near real-time feedback to the operator about the quality of the captured images and whether it is sufficient for downstream analysis, or whether repeat imaging is recommended.

Given the diversity in age and phenotypes of IRDs, Retinograd-AI is a robust starting point for building gradeability models for FAF imaging for other conditions where FAF is commonly used such as Geographic Atrophy and Central Serous Chorioretinopathy, potentially via transfer learning using Retinograd-AI weights as a starting point.

We expect automatic gradability annotations to prove invaluable to future image classification and segmentation tasks as imaging quality is a significant confounder for many image-derived metrics.

In the future, we aim to improve Retinograd-AI by incorporating additional data from other conditions, as well as extend our approach to further imaging modalities.

## Ethics

This research was approved by the IRB and the UK Health Research Authority Research Ethics Committee (REC) reference (22/WA/0049) “Eye2Gene: accelerating the diagnosis of inherited retinal diseases “ Integrated Research Application System (IRAS) (project ID: 242050). All research adhered to the tenets of the Declaration of Helsinki.

## Code availability

The source code for the Retinograd-AI model architecture training and inference is available from https://github.com/Eye2Gene/retinograd-ai.

## Data availability

The data that support the findings of this study are divided into two groups, published data and restricted data. Published data are available from the Github repository. Restricted data are curated for under a UCL Business owned license and cannot be published, to protect patient privacy and intellectual property. Synthetic data derived from the test data has been made available at https://github.com/Eye2Gene/retinograd-ai

## Author contributions

WAW, GN, SAK and IM analysed the data and wrote the manuscript. WAW and NP designed the experiments, analysed data and wrote the manuscript. NP obtained funding. SS, TACG, MDV and SAK analysed the data. All authors have critically reviewed the manuscript.

## Acknowledgement

This work is primarily funded by a NIHR AI Award (AI_AWARD02488) which supported NP, WAW, MM, KB, SD and SM. The research was also supported by a grant from the National Institute for Health Research (NIHR) Biomedical Research Centre (BRC) at Moorfields Eye Hospital NHS Foundation Trust and UCL Institute of Ophthalmology. NP was also previously funded by Moorfields Eye Charity Career Development Award (R190031A). OAM is supported by the Wellcome Trust (206619/Z/17/Z). SA is supported by a scholarship from Qatar National Research Fund (GSRA6-1-0329-19010).This project was also supported by a generous donation by Stephen and Elizabeth Archer in memory of Marion Woods. The hardware used for analysis was supported by the BRC Challenge Fund (BRC3_027). We also gratefully acknowledge the support of NVIDIA Corporation with the donation of the Titan Xp GPU used for this research. The views expressed are those of the authors and not the funding organisations.

## Supplementary

**Supplementary Table 1:** List of hyperparameter settings used for training the neural network.

| Parameter | Value |
| --- | --- |
| Architecture | inception_resnet_v2 |
| Batch size | 4 |
| Image size | (768,768) |
| Train Epochs | 20 (Early stopping as no validation loss improvement) |
| Optimiser | Adam |
| Loss | Weighted categorical cross-entropy |
| Learning rate | 1e-5 |
| Augmentations | Horizontal flipping, Random rotations |

**Supplementary Table 2:** Inter-grader confusion matrix G1/2/3=Grader 1/2/3, G=Gradable, U=Ungradable.

| Grader 1 vs Grader 2 |  | G2 |  | Grader 1 vs Grader 3 |  | G3 |  | Grader 2 vs Grader 3 |  | G3 |  |
| --- | --- | --- | --- | --- | --- | --- | --- | --- | --- | --- | --- |
|  |  | G | U |  |  | G | U |  |  | G | U |
| G1 | G | 110 | 7 | G1 | G | 113 | 4 | G2 | G | 110 | 5 |
|  | U | 5 | 11 |  | U | 2 | 14 |  | U | 5 | 13 |

